## Supplemental Table 1 for "Type 2 diabetes mellitus accelerates brain aging and cognitive decline: complementary findings from UK Biobank and meta-analyses"

| **Supplementary Table 1.** Summary of all relevant UK Biobank data-fields. | | |
| --- | --- | --- |
| **Variable** | **Designation** | **Instance Number** |
| Diagnosis (T2DM) | 2443 | 0, 2 |
| Age | 21003 | 0, 2 |
| Sex | 31 | 2 |
| Education | 6138 | 2 |
| Age of onset (T2DM) | 2976 | 0-2 |
| Blood Pressure | 93, 94, 4079, 4080 | 2 |
| Had Menopause | 2724 | 0-2 |
| Had hormone replacement therapy | 3546 | 2 |
| Body Mass Index (BMI) | 21001 | 2 |
| Medication Status (Metformin) | 20003 | 2 |
| Gray-Matter Volume | 25005-25006, 25782-25920 | 2 |
| Resting-State MRI Images | 20227 | 2 |
| Matrix-Pattern Completion (Abstract Reasoning) | 20016 | 2 |
| Alphanumeric Trail-Making Test (Executive Function) | 6350 | 2 |
| Symbol-Digit Substitution (Processing Speed) | 23324 | 2 |
| Snap Game (Reaction Time) | 20023 | 0, 2 |
| Numeric Memory Test (Numeric Memory) | 4282 | 2 |
