## Supplemental Table 2 for "Type 2 diabetes mellitus accelerates brain aging and cognitive decline: complementary findings from UK Biobank and meta-analyses"

| **Supplementary Table 2.** List and justification for studies excluded from our cognitive meta-analysis. | |
| --- | --- |
| **Studies** | **Justification for Exclusion** |
| Asimakopoulou et al., 2002 | Did not match for education |
| Brands et al., 2007 | Identical sample of study already included |
| Bruehl et al., 2009 | Authors did not provide requested data |
| Callisaya et al., 2018 | Identical sample of study already included |
| Chen et al., 2014 | Inadequate cognitive testing |
| Chen et al., 2017 | Inadequate cognitive testing |
| Christman et al., 2010 | Did not match for education |
| Cooray et al., 2011 | Authors did not provide requested data |
| Cui et al., 2015 | Identical sample of study already included |
| Cui et al., 2017 | Identical sample of study already included |
| Cui et al., 2017 | Identical sample of study already included |
| Degen et al., 2016 | Authors did not provide requested data |
| Dey et al., 1997 | Inadequate cognitive testing |
| Dore et al., 2009 | Authors did not provide requested data |
| Elias et al., 1997 | No baseline data in longitudinal design |
| Grodstein et al., 2001 | Inadequate cognitive testing |
| Hassing et al., 2004 | Unclear sizes of sample sub-groups |
| Helkala et al., 1995 | Did not match for education |
| Kinga & Anett, 2016 | Authors unable to be reached |
| Kumari et al., 2005 | Did not match for education |
| Liu et al., 2016 | Identical sample of study already included |
| Liu et al., 2018 | Identical sample of study already included |
| Liu et al., 2020 | Identical sample of study already included |
| Manschot et al., 2006 | Identical sample of study already included |
| Mooradian et al., 1988 | Inadequate cognitive testing |
| Nazaribadie et al., 2013 | Authors did not provide requested data |
| Nealon et al., 2017 | Did not match for education |
| Nooyens et al., 2010 | Authors did not provide requested data |
| Perlmuter et al., 1984 | Inadequate cognitive testing |
| Ravona-Springer et al., 2018 | Authors did not provide requested data |
| Reijmer et al., 2011 | Identical sample of study already included |
| Robertson-Tchabo et al., 1986 | Inadequate cognitive testing |
| Ruis et al., 2009 | Authors did not provide requested data |
| Scott et al., 1998 | Identical sample of study already included |
| Sinclair et al., 2000 | Inadequate cognitive testing |
| Smith et al., 2009 | Identical sample of study already included |
| Spauwen et al., 2015 | Authors did not provide requested data |
| van Gemert et al., 2018 | Authors did not provide requested data |
| Watari et al., 2006 | Authors did not provide requested data |
| Xia et al., 2013 | Identical sample of study already included |
| Xia et al., 2015 | Identical sample of study already included |
