## Supplemental Table 3 for "Type 2 diabetes mellitus accelerates brain aging and cognitive decline: complementary findings from UK Biobank and meta-analyses"

| **Supplementary Table 3.** Characteristics of patients who underwent cognitive testing in studies included in our meta-analysis. | | | | | | | | |
| --- | --- | --- | --- | --- | --- | --- | --- | --- |
| **Studies** | | ***N*** | | **Age (Mean)** | | **Education (Years)** | | **Female (%)** |
| **Author** | **Year** | **T2DM** | **HC** | **T2DM** | **HC** | **T2DM** | **HC** |  |
| Aberle et al. | 2008 | 38 | 421 | 62.9 | 62.97 | 9.94 | 9.93 | 48.5 |
| Arvanitakis et al. | 2006 | 116 | 766 | 78 | 80.9 | 13.7 | 14.5 | 78 |
| Atiea et al. | 1995 | 40 | 20 | 69.05 | 68.1 | - | - | 31 |
| Bangen et al. | 2015 | 378 | 1115 | 75.4 | 76.3 | 9.9 | 11.2 | 67 |
| Biessels et al. | 2001 | 13 | 16 | 57.7 | 57.9 | 11.2 | 11.4 | 41.3 |
| Brands et al. | 2007 | 119 | 55 | 65.9 | 65.2 | 4* | 4 | 49.4 |
| Cholerton et al. | 2019 | 185 | 261 | 53 | 51.3 | 12 | 12.5 | 70.4 |
| Cosway et al. | 2001 | 33 | 32 | 57.7 | 55.9 | 11.2 | 11.8 | 59.2 |
| Cui et al. | 2014 | 29 | 27 | 58.3 | 57.8 | 10.4 | 10.2 | 55.4 |
| Dai et al. | 2017 | 41 | 32 | 65.51 | 67.28 | 15.35 | 16.05 | 52 |
| Garcia-Casares et al. | 2014 | 25 | 25 | 60 | 57.8 | 18.3 | 18.9 | 38 |
| Kanaya et al. | 2004 | 118 | 632 | 73.55 | 69.2 | - | - | 57.2 |
| Lindeman et al. | 2001 | 188 | 476 | 73.4 | 73.8 | 10.9 | 12.3 | - |
| Liu et al. | 2018 | 32 | 32 | 58.09 | 56.88 | 9 | 12 | 42.2 |
| Lowe et al. | 1994 | 80 | 81 | 59.3 | 55.1 | - | - | 63.9 |
| Mankovsky et al. | 2018 | 93 | 18 | 62.3 | 59.5 | 14.7 | 14.3 | 70.2 |
| Mattei et al. | 2019 | 465 | 711 | 58.9 | 56 | - | - | 73.1 |
| Mehrebian et al. | 2012 | 37 | 22 | 56 | 56 | 14 | 14 | 56.5 |
| Mogi et al. | 2004 | 69 | 27 | 71.6 | 73.4 | 10.4 | 11.4 | 64.5 |
| Moran et al. | 2013 | 350 | 363 | 67.8 | 72.1 | 11.3 | 10.9 | 43.2 |
| Naseer et al. | 2014 | 20 | 20 | 53.3 | - | - | - | - |
| Rawlings et al. | 2015 | 1779 | 11572 | 58.2 | 56.8 | - | - | 55.6 |
| Redondo et al. | 2016 | 20 | 23 | 70.82 | 70.92 | 6.79 | 7.36 | 46 |
| Reijmer et al. | 2016 | 35 | 35 | 71.1 | 71 | 4* | 4 | 41.4 |
| Ryan & Geckle | 2008 | 50 | 50 | 50.8 | 50.5 | 14.4 | 14 | 73 |
| Solanki et al. | 2009 | 50 | 30 | - | - | - | - | - |
| Takeuchi et al. | 2012 | 42 | 32 | 62.4 | 63.8 | 13.7 | 14.5 | 40 |
| van den Berg et al. | 2010 | 68 | 38 | 65.6 | 64.8 | 4* | 4 | 48.1 |
| van Harten et al. | 2007 | 92 | 44 | 73.2 | 72.9 | 4* | 4.4 | 55.8 |
| Xia et al. | 2015 | 38 | 40 | 56 | 57.1 | 9.6 | 10.3 | 51.3 |
| Yau et al. | 2010 | 18 | 18 | 16.46 | 17.16 | 10.75 | 11.15 | - |
| Yeung et al. | 2009 | 41 | 424 | 68.59 | 67.84 | 15.12 | 15.33 | 68 |
| Zhou et al. | 2010 | 21 | 19 | 68 | 69.16 | 12.48 | 13.84 | 50 |
| Zihl et al | 2010 | 12 | 19 | 42.45 | 36.8 | 10.5 | 10.9 | - |
| *T2DM, type-2 diabetes mellitus; HC, healthy control *median education* | | | | | | | | |
