## Supplemental Table 4 for "Type 2 diabetes mellitus accelerates brain aging and cognitive decline: complementary findings from UK Biobank and meta-analyses"

| **Supplementary Table 4.** Summary of cognitive functions assessed, with corresponding instruments. | | |
| --- | --- | --- |
| **Domains** | **Common Tests** | **Description** |
| Abstract Reasoning | Raven’s Progressive Matrices, Matrix Pattern Completion, Weschler Similarities, Standard Progressive Matrices | Manipulation of presented information to solve a problem without prior knowledge. Interrelated with fluid intelligence. Often presented as shape or logic puzzles. |
| Executive Function | Trail Making (B), Stroop (III), Brixton Spatial Anticipation, Wisconsin Card Sort, Color Trails (2), Weschler Letter Number Sequencing | Top-down coordination of other cognitive domains (e.g., memory, motor function) to solve problems and manage cognitive resources. Often exhibited in tasks that require a degree of planning. |
| Processing Speed | Trail Making (A), Digit Symbol Substitution, Stroop (I-II), Choice Reaction Time, Color Trails (1) | Speedy encoding and use of information. Often measured by time-to-completion in tasks that require the manipulation of presented information. |
| Numeric Memory | (Forwards) Digit Span, Digit Vigilance Test | Short-term (~2-3 s) recall of numeric information. |
| Visual Memory | Location Learning, Weschler Visual Memory Subtest, Rey-Osterreith Delayed Recall,  Face Recognition Test | Short and long-term recall of visually encoded information. |
| Verbal Memory | Rey & California Auditory Verbal Learning Tests, Hopkins Verbal Learning Test, Delayed Word Recall, Weschler Text Recall Sub, Word List Recall,  Weschler Story Recall | Short and long-term recall of verbal information. Includes both auditory and visual encoding. |
| Verbal Fluency | Word & Semantic Fluency Tests, Controlled Oral word Association Test, Letter & Category Fluency Tests, Boston Naming Test | Language skills. Commonly measured by enumeration (e.g., name as many words as you can that begin with the letter “B”). |
| Visuospatial Reasoning | Rey-Osterreith Figure Copy, Taylor Complex Figure,  Weschler Object Assembly | Manipulation or reconstruction of spatial information. |
| Working Memory | (Backwards) Digit Span, Corsi Block Tapping, N-back | Holding information for a short time for use on a current task. Characterized by both maintaining and manipulating stored information. Commonly measured by having subjects re-order learned information. |
| *Harvey, (2019). Domains of Cognition and their Assessment. Dialogues of Clinical Neuroscience, 21(3), 227-237. doi:10.31887/DCNS.2019.21.3* | | |
