## Supplemental Table 5 for "Type 2 diabetes mellitus accelerates brain aging and cognitive decline: complementary findings from UK Biobank and meta-analyses"

**Supplementary Table 5.** Studies identified as most relevant for each key word by NeuroQuery algorithm.

|  | T2DM studies | Age studies |
| --- | --- | --- |
| 1. | Chun-Xia Wang et al. 2014 | György A Homola et al. 2012 |
| 2. | Xiangzhe Qiu et al. 2016 | Peiying Liu et al. 2013 |
| 3. | Natalia García-Casares et al. 2016 | G Juckel et al. 2012 |
| 4. | Z-L Wang et al. 2017 | Natalie C Ebner et al. 2013 |
| 5. | Thomas J Marder et al. 2014 | Michelle Hampson et al. 2012 |
| 6. | Franco Cauda et al. 2009 | Sien Hu et al. 2012 |
| 7. | Ying Cui et al. 2015 | Yu-Chien Wu et al. 2011 |
| 8. | Dae-Jin Kim et al. 2016 | Rafat S Mohtasib et al. 2012 |
| 9. | Jung-Lung Hsu et al. 2012 | Vonetta M Dotson et al. 2016 |
| 10. | Zhiye Chen et al. 2012 | Estela Càmara et al. 2007 |
| 11. | Christopher M Marano et al. 2014 | Harri Littow et al. 2010 |
| 12. | Olivia M Farr et al. 2016 | Andrew P Merluzzi et al. 2016 |
| 13. | Dan-Miao Sun et al. 2017 | Emily S Nichols et al. 2016 |
| 14. | Dewang Mao et al. 2015 | Maria Morozova et al. 2016 |
| 15. | Rongfeng Qi et al. 2012 | Kristen M Kennedy et al. 2009 |
| 16. | Dewang Mao et al. 2015 | Chiara Chiapponi et al. 2013 |
| 17. | Xin Huang et al. 2016 | Kathrin Cohen Kadosh et al. 2013 |
| 18. | Wenqing Xia et al. 2013 | Quinton Deeley et al. 2008 |
| 19. | Po Lai Yau et al. 2009 | Kristen M Kennedy et al. 2015 |
| 20. | Reza Tadayonnejad et al. 2019 | Tatia M C Lee et al. 2006 |
| 21. | Chen Liu et al. 2014 | Joshua Carp et al. 2011 |
| 22. | Yue Cheng et al. 2017 | Esther H H Keulers et al. 2010 |
| 23. | Chuanming Li et al. 2014 | Kristin Nordin et al. 2017 |
| 24. | Zhilian Zhao et al. 2014 | Joshua Carp et al. 2010 |
| 25. | Xiaofen Ma et al. 2015 | Mark B Schapiro et al. 2004 |
| 26. | Jessica A Turner et al. 2013 | Nick S Ward et al. 2008 |
| 27. | Jiaxing Zhang et al. 2016 | Nancy E Adleman et al. 2016 |
| 28. | Yingying Yue et al. 2015 | Kaitlin L Bergfield et al. 2010 |
| 29. | Nicola Pannacciulli et al. 2006 | Jenny R Rieck et al. 2017 |
| 30. | Xin Di et al. 2013 | Marco Hirnstein et al. 2011 |
