## Supplemental Table 6 for "Type 2 diabetes mellitus accelerates brain aging and cognitive decline: complementary findings from UK Biobank and meta-analyses"

| **Supplementary Table 6.** Study estimates of cognitive meta-analysis. | | |
| --- | --- | --- |
| **Executive Function** | **d = -0.40, K = 18, *p* < 0.001, Q = 186.9, *I^2^*=88.4%** | |
| **Studies** | **Effect size (d)** | **Confidence Interval (95%)** |
| Bangen et al., 2015 | -0.21 | -0.33, -0.09 |
| Biessels at al., 2001 | -0.66 | -1.41, 0.09 |
| Brands et al., 2007 | -0.54 | -0.86, -0.21 |
| Cui et al., 2014 | -0.54 | -1.07, -0.01 |
| Garcia-Casares et al., 2014 | -0.87 | -1.45, -0.29 |
| Kanaya et al., 2004 | 1.03 | 0.83, 1.24 |
| Lindeman et al., 2001 | -0.09 | -0.26, 0.08 |
| Liu et al., 2018 | -0.54 | -1.04, -0.04 |
| Mehrebian et al., 2012 | -1.35 | -1.93, -0.76 |
| Mogi et al., 2004 | -0.36 | -0.81, 0.08 |
| Reijmer et al., 2016 | -0.33 | -0.80, 0.15 |
| Ryan & Geckle. 2008 | -0.31 | -0.71, 0.08 |
| Takeuchi et al., 2012 | -0.62 | -1.09, -0.15 |
| Van den Berg et al., 2010 | -0.65 | -1.05, -0.24 |
| Xia et al., 2010 | -0.61 | -1.07, -0.16 |
| Yau et al., 2010 | -0.14 | -0.80, 0.51 |
| Yeung et al., 2009 | -0.50 | -0.83, -0.18 |
| Zhou et al., 2010 | -0.48 | -1.10, 0.15 |
| **Immediate Verbal Memory** | **d= –0.39, K=23, *p*<0.001, Q=143.6, *I^2^*= 91.1%** | |
| **Studies** | **Effect size (d)** | **Confidence Interval (95%)** |
| Aberle et al., 2008 | 0.02 | -0.31, 0.36 |
| Arvanitakis et al., 2006 | -0.05 | -0.25, 0.14 |
| Bangen et al., 2015 | -0.13 | -0.25, -0.02 |
| Brands et al., 2007 | -0.32 | -0.64, 0.00 |
| Cholerton et al., 2019 | 0.06 | -0.13, 0.25 |
| Cosway et al., 2001 | -0.32 | -0.81, 0.17 |
| Cui et al., 2014 | -0.07 | -0.59, 0.46 |
| Dai et al., 2017 | -1.7 | -2.24, -1.16 |
| Garcia-Casares et al., 2014 | -1.62 | -2.26, -0.98 |
| Liu et al., 2018 | -0.49 | -0.99, 0.01 |
| Lowe et al., 1994 | -0.06 | -0.36, 0.25 |
| Mattei et al., 2019 | -0.3 | -0.42, -0.19 |
| Mehrebian et al., 2012 | -1.71 | -2.33, -1.10 |
| Mogi et al., 2004 | -0.29 | -0.74, 0.16 |
| Moran et al., 2013 | 0.31 | 0.16, 0.46 |
| Reijmer et al., 2016 | -0.23 | -0.70, 0.24 |
| Ryan & Geckle, 2008 | -0.41 | -0.81, -0.01 |
| Takeuchi et al., 2012 | -0.56 | -1.03, -0.09 |
| van den Berg et al., 2010 | -0.39 | -0.79, 0.01 |
| van Harten et al., 2007 | -0.35 | -0.71, 0.01 |
| Xia et al, 2015 | -0.32 | -0.77, 0.13 |
| Yau et al., 2010 | -0.70 | -1.37, -0.03 |
| Yeung et al., 2009 | -0.37 | -0.69, -0.04 |
| **Verbal Fluency** | **d= –0.37, K=25, *p*<0.001, Q=101.3, *I^2^*=83.3%** | |
| **Studies** | **Effect Size (d)** | **Confidence Interval (95%)** |
| Aberle et al., 2008 | 0.06 | -0.27, 0.39 |
| Arvanitakis et al., 2006 | -0.12 | -0.31, 0.08 |
| Atiea et al., 1995 | -0.32 | -0.86, 0.22 |
| Bangen et al., 2015 | -0.33 | -0.44, -0.21 |
| Brands et al., 2007 | -0.50 | -0.82, -0.17 |
| Cholerton et al., 2019 | -0.37 | -0.56, -0.18 |
| Cosway et al., 2001 | -0.30 | -0.79, 0.19 |
| Dai et al., 2017 | -0.83 | -1.31, -0.35 |
| Garcia-Casares et al., 2014 | -0.92 | -1.5, -0.34 |
| Kanaya et al., 2004 | -1.10 | -1.31, -0.9 |
| Liu et al., 2018 | -0.44 | -0.94, 0.05 |
| Lowe et al., 1994 | -0.46 | -0.77, -0.15 |
| Mankovsky et al., 2018 | -0.32 | -0.83, 0.19 |
| Mattei et al., 2019 | -0.33 | -0.44, -0.21 |
| Mehrebian et al., 2012 | -0.18 | -0.71, 0.34 |
| Moran et al., 2013 | -0.03 | -0.18, 0.12 |
| Rawlings et al., 2015 | -0.36 | -0.41, -0.31 |
| Reijmer et al., 2016 | -0.15 | -0.62, 0.32 |
| Solanki et al., 2009 | -0.47 | -0.93, -0.01 |
| Takeuchi et al., 2012 | -0.59 | -1.06, -0.12 |
| van Harten et al., 2007 | -0.64 | -1.01, -0.27 |
| Yau et al., 2010 | -0.07 | -0.72, 0.59 |
| Yeung et al., 2009 | -0.15 | -0.47, 0.17 |
| Zhou et al., 2010 | -0.58 | -1.21, 0.06 |
| Zihl et al., 2010 | 0.26 | -0.46, 0.99 |
| **Working Memory** | **d= –0.36, K=12, *p*<0.001, Q=30.2, *I^2^*=72.2%** | |
| **Studies** | **Effect Size (d)** | **Confidence Interval (95%)** |
| Arvanitakis et al., 2006 | -0.04 | -0.24, 0.15 |
| Atiea et al., 1995 | -0.48 | -1.02, 0.07 |
| Biessels et al., 2001 | -0.63 | -1.38, 0.12 |
| Brands et al., 2007 | -0.36 | -0.69, -0.04 |
| Lowe et al., 1994 | -0.04 | -0.35, 0.26 |
| Mankovsky et al., 2018 | -0.07 | -0.57, 0.44 |
| Mattei et al., 2019 | -0.27 | -0.38, -0.15 |
| Ryan & Geckle, 2008 | -0.34 | -0.73, 0.06 |
| Solanki et al., 2009 | -1.33 | -1.83, -0.84 |
| Takeuchi et al, 2012 | -0.56 | -1.03, -0.10 |
| van den Berg et al., 2010 | -0.50 | -0.90, -0.10 |
| Yau et al., 2010 | -0.17 | -0.82, 0.49 |
| **Abstract Reasoning** | **d= –0.36, K=8, *p*<0.001, Q=10.57, *I^2^*=29.2%** | |
| **Studies** | **Effect Size (d)** | **Confidence Interval (95%)** |
| Arvanitakis et al., 2006 | -0.24 | -0.43, -0.04 |
| Bangen et al., 2015 | -0.48 | -0.59, -0.36 |
| Brands et al., 2007 | -0.19 | -0.51, 0.13 |
| Cosway et al., 2001 | -0.10 | -0.59, 0.38 |
| Lowe et al., 1994 | -0.41 | -0.73, -0.10 |
| Ryan & Geckle, 2008 | -0.27 | -0.67, 0.12 |
| van den Berg et al., 2010 | -0.38 | -0.78, 0.02 |
| Zihl et al., 2010 | -1.06 | -1.82, -0.29 |
| **Processing Speed** | **d= –0.34, K=31, *p*<0.001, Q=227.3, *I^2^*=82.3 %** | |
| **Studies** | **Effect Size (d)** | **Confidence Interval (95%)** |
| Aberle et al., 2008 | 0.00 | -0.33, 0.33 |
| Arvanitakis et al., 2006 | -0.14 | -0.34, 0.05 |
| Atiea et al., 1995 | 0.05 | -0.49, 0.59 |
| Bangen et al., 2015 | -0.22 | -0.34, -0.11 |
| Biessels et al., 2001 | 0.19 | -0.54, 0.92 |
| Brands et al., 2007 | -0.26 | -0.58, 0.06 |
| Cholerton et al., 2019 | -0.43 | -0.62, -0.24 |
| Cosway et al., 2001 | -0.30 | -0.79, 0.19 |
| Cui et al., 2014 | -0.68 | -1.22, -0.14 |
| Dai et al., 2017 | 0.25 | -0.21, 0.72 |
| Garcia-Casares et al., 2014 | -0.66 | -1.23, -0.09 |
| Lindeman et al., 2001 | -0.02 | -0.19, 0.15 |
| Liu et al., 2018 | -0.36 | -0.86, 0.13 |
| Mattei et al., 2019 | -0.37 | -0.49, -0.25 |
| Mehrebian et al., 2012 | -1.23 | -1.8, -0.65 |
| Mogi et al., 2004 | -0.59 | -1.04, -0.14 |
| Moran et al., 2013 | 0.16 | 0.02, 0.31 |
| Naseer et al., 2014 | -0.61 | -1.25, 0.02 |
| Rawlings et al., 2015 | -0.65 | -0.70, -0.60 |
| Redondo et al., 2016 | -0.54 | -1.15, 0.07 |
| Reijmer et al., 2016 | -0.06 | -0.53, 0.41 |
| Ryan & Geckle, 2008 | -0.43 | -0.83, -0.03 |
| Solanki et al., 2009 | -0.93 | -1.41, -0.46 |
| Takeuchi et al., 2012 | -0.61 | -1.08, -0.14 |
| van den Berg et al., 2010 | -0.11 | -0.51, 0.29 |
| van Harten et al., 2007 | -0.48 | -0.84, -0.11 |
| Xia et al, 2015 | -0.27 | -0.71, 0.18 |
| Yau et al., 2010 | -0.69 | -1.37, -0.02 |
| Yeung et al., 2009 | -0.38 | -0.70, -0.06 |
| Zhou et al., 2010 | -0.27 | -0.89, 0.35 |
| Zihl et al., 2010 | -1.28 | -2.07, -0.49 |
| **Visuospatial Reasoning** | **d= –0.32, K=13, *p*<0.001, Q=27.3, *I^2^*=56.6%** | |
| **Studies** | **Effect Size (d)** | **Confidence Interval (95%)** |
| Arvanitakis et al., 2006 | -0.11 | -0.31, 0.09 |
| Bangen et al., 2015 | -0.18 | -0.30, -0.07 |
| Biessels et al., 2001 | -0.25 | -0.98, 0.49 |
| Brands et al., 2007 | -0.21 | -0.53, 0.11 |
| Garcia-Casares et al., 2014 | -0.79 | -1.36, -0.21 |
| Lowe et al., 1994 | -0.16 | -0.47, 0.15 |
| Mattei et al., 2019 | -0.33 | -0.45, -0.21 |
| Moran et al., 2013 | -0.56 | -0.71, -0.41 |
| Ryan & Geckle, 2008 | -0.38 | -0.78, 0.01 |
| Takeuchi et al., 2012 | -0.41 | -0.87, 0.06 |
| van den Berg et al., 2010 | -0.16 | -0.56, 0.24 |
| Xia et al, 2015 | -0.71 | -1.17, -0.25 |
| Zhou et al., 2010 | -0.47 | -1.10, 0.16 |
| **Delayed Verbal Memory** | **d= –0.21, *p*<0.001, K=21, Q=114.3, *I^2^*=77.5%** | |
| **Studies** | **Effect Size (d)** | **Confidence Interval (95%)** |
| Arvanitakis et al., 2006 | -0.01 | -0.21, 0.18 |
| Bangen et al., 2015 | -0.07 | -0.19, 0.05 |
| Brands et al., 2007 | -0.28 | -0.60, 0.04 |
| Cholerton et al., 2019 | 0.04 | -0.15, 0.23 |
| Cosway et al., 2001 | -0.24 | -0.73, 0.24 |
| Cui et al., 2014 | -0.08 | -0.60, 0.45 |
| Dai et al., 2017 | -0.75 | -1.22, -0.27 |
| Liu et al., 2018 | -0.62 | -1.13, -0.12 |
| Lowe et al., 1994 | -0.10 | -0.41, 0.21 |
| Mehrebian et al., 2012 | -0.76 | -1.31, -0.22 |
| Mogi et al., 2004 | 0.18 | -0.26, 0.63 |
| Moran et al., 2013 | 0.20 | 0.05, 0.35 |
| Rawlings et al., 2015 | -0.40 | -0.45, -0.35 |
| Reijmer et al., 2016 | -0.13 | -0.60, 0.34 |
| Ryan & Geckle, 2008 | -0.22 | -0.61, 0.17 |
| Takeuchi et al., 2012 | -0.36 | -0.82, 0.10 |
| van den Berg et al., 2010 | -0.40 | -0.80, 0.00 |
| van Harten et al., 2007 | -0.42 | -0.78, -0.05 |
| Xia et al, 2015 | -0.15 | -0.59, 0.30 |
| Yeung et al., 2009 | -0.07 | -0.39, 0.25 |
| Zhou et al., 2010 | -0.96 | -1.62, -0.31 |
| **Numeric Memory** | **d= –0.21, *p*=0.005, K=16, Q=38.1, *I^2^*=70.2%** | |
| **Studies** | **Effect Size (d)** | **Confidence Interval (95%)** |
| Arvanitakis et al., 2006 | -0.13 | -0.33, 0.06 |
| Atiea et al., 1995 | -0.47 | -1.01, 0.08 |
| Biessels et al., 2001 | -1.05 | -1.83, -0.27 |
| Brands et al., 2007 | -0.08 | -0.40, 0.24 |
| Dai et al., 2017 | -0.66 | -1.14, -0.19 |
| Lindeman et al., 2001 | -0.17 | -0.33, 0.00 |
| Liu et al., 2018 | -0.53 | -1.03, -0.03 |
| Lowe et al., 1994 | 0.14 | -0.17, 0.45 |
| Mankovsky et al., 2018 | 0.17 | -0.34, 0.67 |
| Mattei et al., 2019 | -0.11 | -0.22, 0.01 |
| Moran et al., 2013 | 0.08 | -0.07, 0.22 |
| Naseer et al., 2014 | 0.04 | -0.57, 0.66 |
| Solanki et al., 2009 | -0.92 | -1.39, -0.44 |
| Takeuchi et al., 2012 | -0.33 | -0.80, 0.13 |
| van den Berg et al., 2010 | -0.12 | -0.52, 0.27 |
| Yau et al., 2010 | -0.30 | -0.96, 0.36 |
| **Recognition Verbal Memory** | **d= –0.21, *p*=0.01, K=12, Q=37.4, *I^2^*=78.8%** | |
| **Studies** | **Effect Size (d)** | **Confidence Interval (95%)** |
| Arvanitakis et al., 2006 | 0.00 | -0.20, 0.20 |
| Bangen et al., 2015 | -0.06 | -0.17, 0.06 |
| Brands et al., 2007 | -0.38 | -0.70, -0.06 |
| Dai et al., 2017 | -0.61 | -1.08, -0.14 |
| Liu et al., 2018 | -0.53 | -1.02, -0.03 |
| Lowe et al., 1994 | -0.07 | -0.38, 0.24 |
| Mattei et al., 2019 | -0.17 | -0.28, -0.05 |
| Mehrebian et al., 2012 | -0.46 | -1.00, 0.07 |
| Moran et al., 2013 | 0.16 | 0.01, 0.31 |
| Takeuchi et al., 2012 | 0.12 | -0.34, 0.58 |
| van den Berg et al., 2010 | -0.51 | -0.92, -0.11 |
| Zhou et al., 2010 | -0.89 | -1.55, -0.24 |
| **Visual Memory** | **d= –0.13, *p*=0.32, K=8, Q=19.9, *I^2^*=66.6%** | |
| **Studies** | **Effect Size (d)** | **Confidence Interval (95%)** |
| Aberle et al., 2008 | -0.16 | -0.49, 0.18 |
| Brands et al., 2007 | 0.19 | -0.13, 0.51 |
| Cosway et al., 2001 | -0.10 | -0.59, 0.39 |
| Lowe et al., 1994 | 0.20 | -0.11, 0.51 |
| Solanki et al., 2009 | -0.96 | -1.43, -0.48 |
| Takeuchi et al., 2012 | -0.27 | -0.73, 0.19 |
| van den Berg et al., 2010 | 0.00 | -0.39, 0.40 |
| Yau et al., 2010 | -0.13 | -0.79, 0.52 |
